# Genomic and Transcriptomic Correlates of Response to Tarlatamab in Small Cell Lung Cancer

**DOI:** 10.64898/2026.01.30.26344966

**Authors:** Zofia Cieslak, Drew T. Bergman, Donald C. Green, Rutu S. Vyas, Aidan Lackstrom, Shawn M. Balcome, Kyle J. Syme, Nidhi Shah, Ivy Riano, Laura J. Tafe, Xiaoying Liu, Mehmet K. Samur, Louis J. Vaickus, Konstantin H. Dragnev, Alexander D. Fuld, Keisuke Shirai, Parth S. Shah

**Affiliations:** Geisel School of Medicine at Dartmouth; Department of Radiation Oncology, Memorial Sloan Kettering Cancer Center; Department of Radiation Oncology and Applied Sciences, Dartmouth Cancer Center; Department of Pathology and Laboratory Medicine, Dartmouth-Hitchcock Medical Center; Boston University; Department of Genetics and Child Development, Dartmouth Health Children’s; Department of Hematology & Oncology, Dartmouth Cancer Center; Department of Data Science, Dana Farber Cancer Institute

## Abstract

**Purpose:** Tarlatamab is a DLL3-directed bispecific T-cell engager demonstrating clinically meaningful activity in relapsed small cell lung cancer (SCLC) in the phase II DeLLphi-301 trial. Determinants of tarlatamab sensitivity and resistance are incompletely understood, and thus we sought to identify genomic and transcriptional correlates of tarlatamab sensitivity using a clinical sequencing pipeline at a single comprehensive cancer center.

**Experimental Design:** We performed a retrospective, single-institution analysis of 12 patients with SCLC treated with tarlatamab. Whole-exome sequencing (WES) and exome-capture whole-transcriptome sequencing (WTS) were performed on 12 samples, and two matched samples after treatment with tarlatamab. Integrative analysis examined correlation between molecular features and clinical outcomes.

**Results:** The overall response rate was 50%, which was consistent with outcomes reported in the DeLLphi-301 trial. Differences between SCLC driver alterations and tumor mutational burden were not significant between responders and non-responders, but homologous recombination deficiency scores were higher in responsive tumors. *DLL3* expression was significantly greater in responders and demonstrated predictive discrimination for clinical response (AUC 0.83). Tumors responsive to tarlatamab were predominantly ASCL1-driven (SCLC-A) and demonstrated increased immune activation, such as enrichment of cytotoxic T-cell, NK-cell, and T cell transcriptional programs. Transcriptional subtype and a composite metric consisting of *DLL3* expression and immune activity (DLI score) further discriminated between responders and non-responders (sensitivity 0.83, specificity 1). Paired post-treatment sample analysis identified loss of ASCL1 lineage and emergence of *YAP1* expression and downregulation of *DLL3*, consistent with lineage plasticity as a mechanism of acquired resistance.

**Conclusions:** Sensitivity to tarlatamab is correlated with a combination of increased *DLL3* expression, ASCL1-driven lineage, and an increased immune activation. Lineage state reprogramming and decrease in *DLL3* expression accompany acquired resistance to tarlatamab. These findings highlight the utility of RNA based biomarkers which integrate target expression, lineage state, and immune context to guide tarlatamab therapy in SCLC. Prospective validation of the whole-transcriptome DLI score and transcriptional subtype will inform tarlatamab response prediction.

## Introduction

Small-cell lung cancer (SCLC) is an aggressive neuroendocrine malignancy with early metastatic spread, rapid progression, and poor survival outcomes^1^ despite the use of immune checkpoint inhibitors (ICI)^2^ for extensive stage disease and advances in next line therapy^3^.

Molecular profiling of SCLC has revealed profound heterogeneity among tumors. Transcription factors define transcriptional subtypes such as ASCL1, NEUROD1, and POU2F3^4^ and their interaction with the microenvironment, influencing response to ICI^5^. Heterogeneity among SCLC subtype and response emphasizes the importance of matching appropriate therapy and exploiting subtype switching to extend response duration.

DLL3, a Notch ligand that is aberrantly expressed in SCLC, serves as the target for tarlatamab, a bispecific T-cell engager that has demonstrated encouraging efficacy and tolerable safety in trials compared with historical outcomes for relapsed disease^6–8^. However, the determinants of sensitivity and resistance to tarlatamab remain unknown. Although *DLL3* expression varies by molecular subtype and tarlatamab relies on CD3-mediated T-cell engagement^9^, the extent to which lineage state and immune activity influence clinical response has not yet been systematically evaluated.

Here, we present an integrated whole-exome (WES) and exome-capture whole-transcriptome (WTS) analysis of tarlatamab treated SCLC to investigate molecular correlates of response and resistance. We profiled 12 consecutive patients treated at a single academic cancer center, including 2 matched post-progression samples, and contextualized these data within a large cohort of 307 publicly available SCLC transcriptomes. This multi-omic framework enables exploration of how SCLC lineage state, genomic architecture, and tumor immune context may converge to influence the biology of DLL3-targeted therapy and provides a foundation for future biomarker development of this highly plastic and molecularly heterogeneous cancer.

## Methods

### Study Design and Patients

This is a retrospective study assessing patients with SCLC who were diagnosed with biopsy and received tarlatamab between July 2024 and November 2025 at a single cancer center. Patient eligibility was determined by a history of platinum-based chemotherapy and at least two tarlatamab infusions, measurable disease according to Response Evaluation Criteria in Solid Tumors (RECIST) version 1.1^10^, and available tumor tissue. Patient characteristic, treatment, and clinical course data were gathered through chart review. The study protocol was approved by the institutional review board.

### Treatment and Response Assessment

Step-up dosing of tarlatamab consisted of the following regimen: 1 mg on day 1, 10 mg on days 8 and 15, and subsequently 10 mg every 2 weeks until disease progression or unacceptable toxicity. The first two infusions, on days 1 and 8, were performed in the inpatient setting to assess for adverse effects such as cytokine release syndrome (CRS) and immune effector cell associated neurotoxicity syndrome (ICANS). Imaging at baseline was defined as the most recent CT or PET-CT obtained prior to or within 1 day of the first tarlatamab infusion, and tumor response to tarlatamab was assessed using RECIST 1.1.

### Tumor Immunohistochemistry and Morphology

Tumor characteristics including synaptophysin, TTF1, Ki67 (12/12 patients) and RB1 (11/12 patients) were obtained through immunohistochemistry staining. Tumor infiltrating lymphocyte (TIL) counts and ratios were obtained for 11/12 patients. H&E-stained whole slide images (WSI) were created for each biopsy and used for NGS analysis. The images were loaded into QuPath (https://qupath.github.io/) for annotation. For each WSI, three representative regions were chosen and designated using a rectangle annotation. Within each rectangle, lymphocytes were annotated using the dot annotation tool. TIL count ratio was calculated as the number of total TILs across all three regions divided by the area in pixels of all three annotation regions (TIL / annotation area in pixels).

### Whole-Exome Sequencing

FFPE DNA was extracted using the Purigen Ionic FFPE to Pure DNA Kit (Purigen Biosystems). DNA quantity was measured using the Qubit dsDNA High Sensitivity assay (ThermoFisher Scientific). Libraries were prepared with the SureSelect XTHS kit and V8 probe set (Agilent) on the Magnis automated platform, with a no-template control included on each run. Final libraries were assessed on the Agilent 4150 TapeStation and sequenced in batches of up to 64 samples on the Illumina NovaSeq 6000 or 6 libraries on the Element AVITI^TM^. A customized implementation of AUGMET v15 (Basesolve Informatics) served as the bioinformatics pipeline for demultiplexing, alignment, variant calling, and QC^11^. The pipeline includes SNP/indel calling, CNV detection for events ≥5 copies, visualization tools, and assay QC metrics; copy-neutral LOH and CNVs <5 copies were not evaluated. SNVs/indels were filtered for sequencing depth (DP ≥ 50), allele frequency (AF ≥ 0.05 with ≥5 alternate reads), and germline frequency (gnomAD genome AF < 0.001 or missing). Variants were restricted to functional classes (missense, nonsense, frameshift, in-frame, splice) and to a predefined SCLC driver gene list for alteration summary^12^. CNVs were classified using log2 copy-ratio thresholds: deep deletion (<–1.2), heterozygous loss (–1.2 to –0.6), diploid (–0.6 to 0.4), gain (0.4 to 0.75), and amplification (>0.75). Only genes meeting these defined categories were retained for downstream analysis.

### Whole-Transcriptome Sequencing

Sample preparation was performed as previously described^13^. Briefly, RNA was extracted from FFPE samples using the Purigen Ionic FFPE to Pure RNA Kit (Purigen Biosystems, Pleasanton, CA) and quantified using the Qubit RNA Quantitation, High Sensitivity kit (Thermo Fisher Scientific, Waltham, MA). Library preparation was performed using the Illumina RNA Prep with Enrichment Tagmentation kit and Illumina Exome Panel (Illumina, San Diego, CA) on 100 ng of RNA. Samples were indexed with Illumina DNA/RNA UD Indexes (Illumina 20027213), and final libraries were quality checked using the Agilent 4150 TapeStation. Libraries were sequenced on the Illumina NovaSeq 600. Demultiplexing, read alignment, and counting is performed by the automated bioinformatics pipeline AUGMET^11^. Reads were pseudoaligned to the GRCh37 reference transcriptome to generate transcript-level abundance estimates^14^. Transcript-level estimates were then imported into R using tximport^15^ and summarized to gene-level counts.

### Supplemental SCLC Whole-Transcriptome Data

Publicly available SCLC transcriptomic datasets were incorporated to enable cross-cohort comparison and molecular subtype analysis. Processed RNA-seq count matrices were downloaded from two sources: (1) a 226-sample SCLC cohort from https://github.com/SeHoonLab/molecular-subtype-SCLC^16^, and (2) 86 sample SCLC cohort from GEO accession GSE60052^17^. Count matrices were merged with our processed gene-level counts and restricted to shared genes. Sample metadata were harmonized and annotated with batch labels corresponding to the three contributing sources. Batch correction was performed using ComBat (sva^18^), modeling batch as the only covariate and allowing expression values to be adjusted across datasets. Genes with all-missing values, non-finite values, or zero variance after correction were removed. The resulting ComBat-adjusted matrix served as the integrated dataset for all downstream analyses, including subtype classification and dimensionality reduction.

### Transcriptional Subtype Classification and Gene Signature Analysis

Transcriptional subtypes were assigned using a supervised approach (adapted from Park et al.^16^) based on z-normalized expression signature scores for ASCL1^19^, NEUROD1^19^, and POU2F3/Tuft programs^20^. Signature scores were computed using the singscore framework^21^, which ranks genes within each sample and calculates normalized mean ranks for each signature. These normalized singscore values were used directly for subtype calling. Subtypes were then assigned using a hierarchical classifier: SCLC-P for tuft-high tumors (>90th percentile) with ASCL1 and NEUROD1 both below their medians; SCLC-TN for tumors with ASCL1 <32nd percentile and NEUROD1 <35th percentile; SCLC-N for NEUROD1 above its median with ASCL1 <25th percentile; SCLC-A for ASCL1 ≥32nd percentile with NEUROD1 <median; and SCLC-AN for intermediate or co-expressed profiles not meeting the above criteria. Additional signatures including neuroendocrine and non-neuroendocrine^22^, ASCL1/NEUROD1 shared targets^19^, MYC-related Notch^22^, NK cell^23^, cytolytic T^24^, and T-cell–inflamed^25^ programs were quantified for downstream analyses. A tumor immune activity score was calculated as the mean of the NK-cell, cytolytic T-cell, and T-cell–inflamed signature scores, providing a single effector-immune metric across samples. Finally, a composite *DLL3* and immune activity (DLI) score was calculated as the sum of the scaled mean *DLL3* TPM and immune activity score.

### Bioinformatics Analysis and Statistical Methods

Analyses were performed in R (v4.5.1). RNA-seq count data were filtered to protein-coding, sufficiently expressed genes and normalized using DESeq2^26^. Differential expression was estimated with DESeq2 Wald tests and log2 fold-change shrinkage with apeglm^27^. Gene set enrichment analysis was performed using clusterProfiler^28^. Transcription factor and immune signature scores were calculated using the singscore framework as above^21^. Predictive performance of *DLL3* expression was evaluated with ROC analysis and Youden’s J statistic^21,29^. Heatmaps were generated using ComplexHeatmap^30^, and visualizations using ggplot2^31^. Schematic figures were created with BioRender.

### Reproducibility and Data Availability

Clinical data summaries and exome sequencing summary metrics are provided in the supplemental materials. Additional processed data and analytic code are available from the corresponding author upon reasonable request and pending institutional approval.

## Results

### Patient Characteristics

Twelve consecutive patients with SCLC treated with tarlatamab at a single cancer center were included, most of whom were current or former heavy smokers and had received prior systemic therapy. Baseline disease was mostly extensive stage, and sites commonly involved the liver, bone, or adrenal glands, and several patients had treated brain metastases or were receiving corticosteroids at treatment initiation. Tumor specimens used for molecular profiling were primarily from primary lung and nodal sites, with a subset from metastatic lesions, and two patients had matched post-treatment biopsies available for analysis (Table 1).

**Table 1.** Patient and Sample Characteristics.

| Characteristic | Tarlatamab<br>(N = 12) |
| --- | --- |
| Sex – no. (%) |  |
| Male | 4 (33) |
| Female | 8 (67) |
| Median age (range) – yr | 66 (47-82) |
| Smoking history – no. (%) |  |
| Non-smoker | 0 (0) |
| Smoker | 12 (100) |
| Median previous lines of therapy (range) | 2 (1-3) |
| Previous PD-L1 antibody therapy– no. (%) |  |
| Yes | 8 (67) |
| No | 4 (33) |
| Brain metastases – no. (%) |  |
| Yes | 7 (58) |
| No | 5 (42) |
| Liver metastases – no. (%) |  |
| Yes | 6 (50) |
| No | 6 (50) |
| Biopsy site |  |
| Primary | 4 (33) |
| Lymph node | 6 (50) |
| Metastasis | 2 (17) |

Cytokine release syndrome occurred in 25% of patients (grade ≥3 in 0%), and ICANS in 25%. The overall response rate was 50% (6/12), disease control rate was 58.3% (7/12). Both the median progression free survival (4.60 vs 1.64 months) and overall survival (10.84 vs 5.90 months) were longer in responders compared to non-responders. At time of cutoff, 3/12 patients (25%) had ongoing responses (Figure 1B).

**Figure 1.**
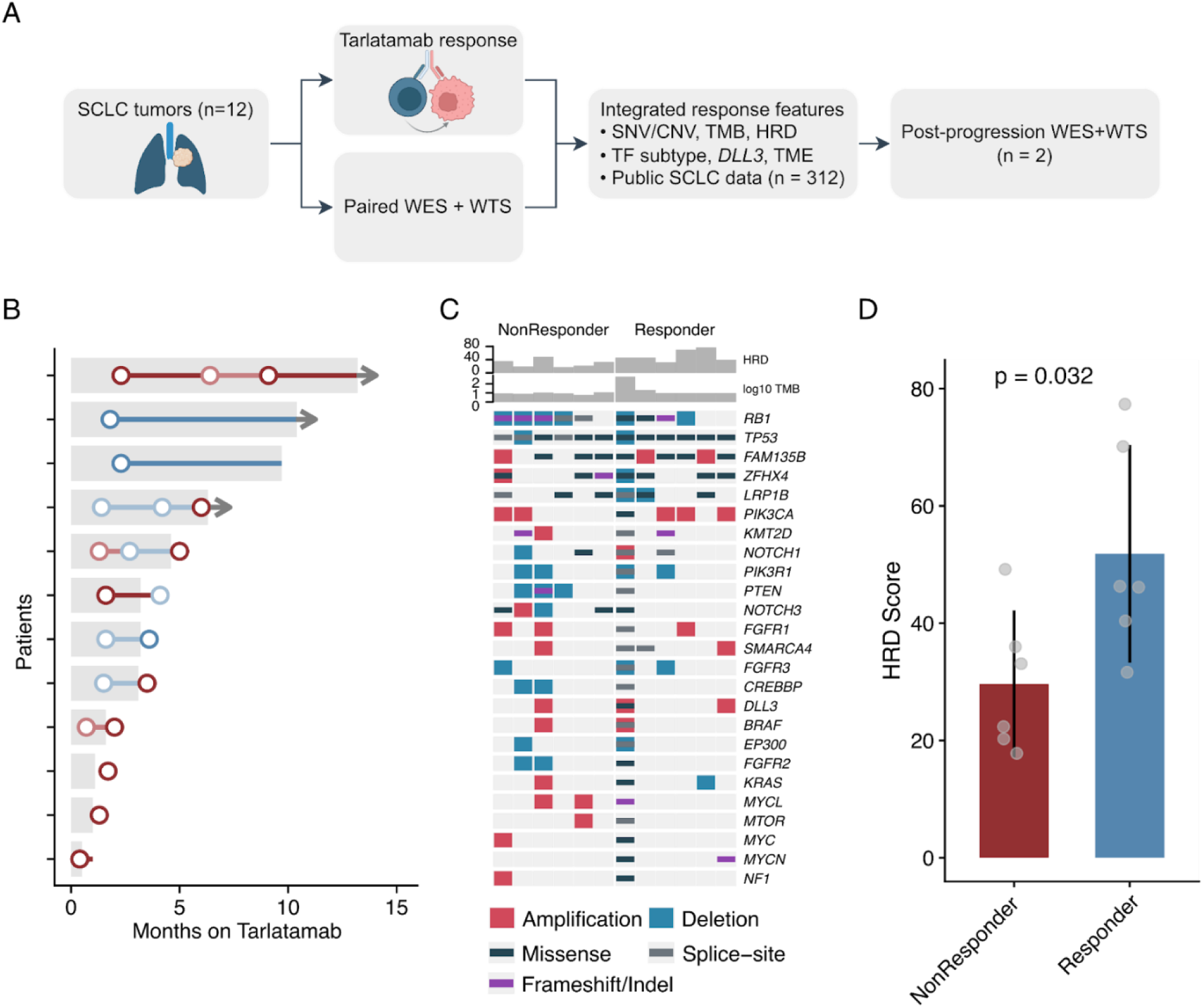
(A) Study outline. Created with use of BioRender. (B) Swimmer’s plot describing SCLC patients’ duration of and response to (light blue = partial response, blue = complete response, light red = stable disease, red = progression) Tarlatamab therapy. Arrows denote ongoing Tarlatamab therapy at cutoff. (C) Oncoprint of key SCLC driver genes by Tarlatamab response by responder status with homologous recombination deficiency (HRD) score and log_10_ tumor mutational burden (TMB). (D) HRD score by Tarlatamab response (p-value, t-test, error bar 95% CI).

### Sample Characteristics

Immunohistochemistry staining was performed for 12 pre-treatment and 2 post-treatment samples. 12/12 tumors were synaptophysin positive, 10/11 were TTF1 positive, 10/10 were RB1 negative, and the Ki67 range was 70-95%.

### Genomic Landscape of SCLC Tarlatamab Response

We next characterized the genomic and transcriptomic landscape of the SCLC tumors prior to tarlatamab treatment using an integrated WES and WTS assay (DH-CancerSeq). All samples passed quality control, yielding high-depth WES (median on-target coverage 277.6x, table S1) and high-quality RNA-seq data (table S1). We characterized the landscape of genomic alterations in our cohort including single nucleotide (SNV) and copy number variants (CNV) from WES and overall did not find strong associations with response to tarlatamab. For example, we identified SNVs and CNVs in key recurrent driver genes of SCLC including *TP53* (12/12), and *RB* (9/12, Fig 1C), which was in line with RB1 loss seen with immunohistochemistry. Copy number analysis also showed copy number variants in key recurrent drivers such as *PIK3CA* amplification, *MYC* family amplification, and *MTOR* amplification (Fig 1C). Although there were modest differences in the number of alterations in the genes described above, there were no significant differences between responders and non-responders given the limited sample size (Fig. S1).

We examined key signatures of overall tumor mutational burden (TMB) and homologous recombination deficiency (HRD) as potential proxies for increased genomic instability, chronic immune activation and signaling, and increased susceptibility to immune effector-mediated killing. TMB did not differ between response groups (median TMB 10 in responders vs 10 in non-responders, p = 0.121, Mann–Whitney U test), except for one hypermutated responder (640 mut/Mb, Figure 1C). HRD scores were significantly higher in the responder group (mean 51.8 vs 29.7, p = 0.032, t-test, Figure 1D).

These findings indicate that while common driver alterations were not predictive of response, higher HRD scores in responders raise the possibility that defects in DNA repair and genomic instability may modulate sensitivity to tarlatamab.

### Higher *DLL3* expression is associated with response to tarlatamab

We next examined whole-transcriptome data to identify transcriptional correlates of tarlatamab sensitivity. In the phase 2 DeLLphi-301 trial, DLL3 was assessed using a low IHC cutoff and responses occurred in both DLL3-positive and DLL3-negative tumors, leaving the role of *DLL3* gene expression unclear^8^. We therefore evaluated whether *DLL3* transcript levels correlated with treatment outcome and found that expression was significantly higher in responders than in non-responders (mean 4.53 vs 2.43 log₂ TPM; p = 0.028, t-test, Figure 2A).

**Figure 2.**
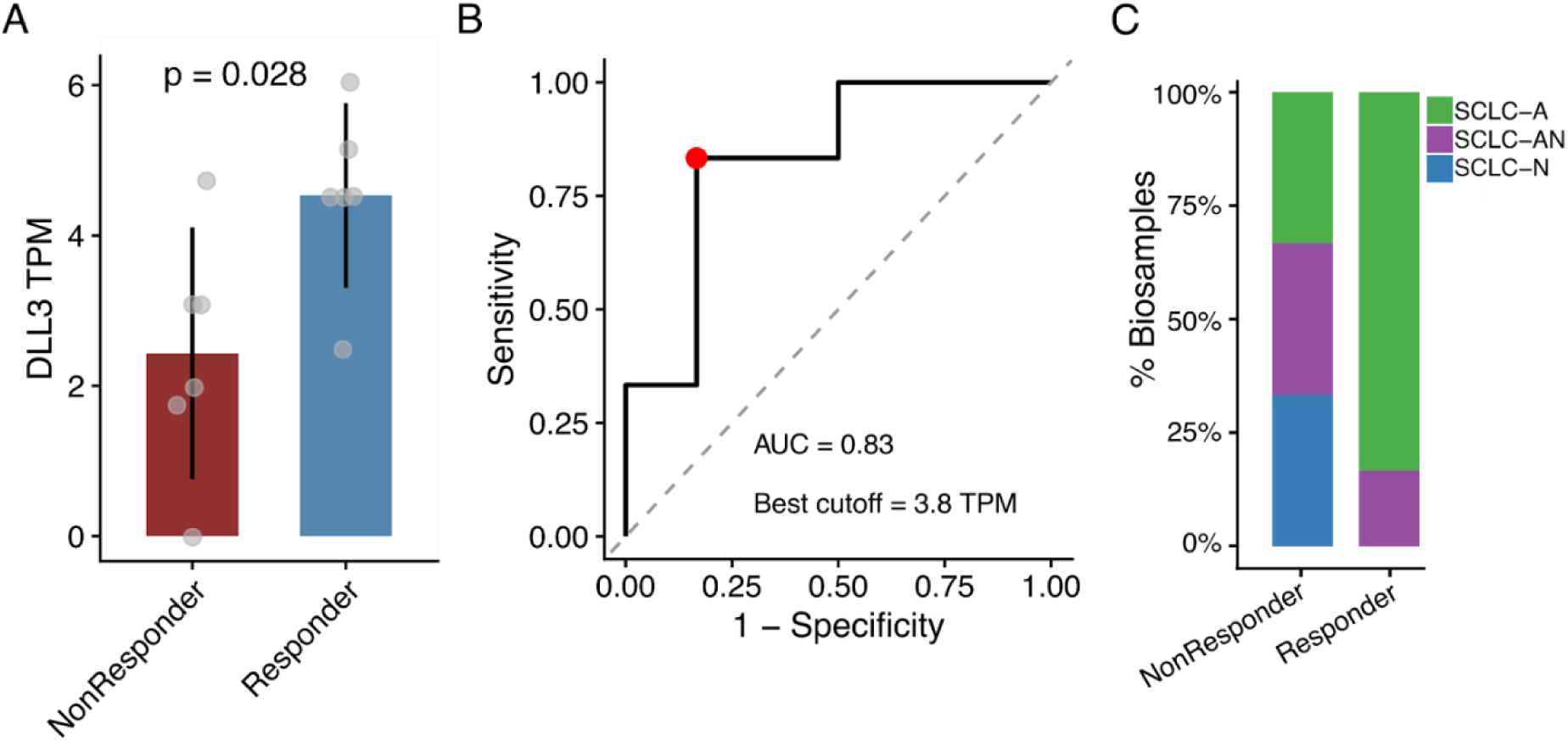
(A) DLL3 expression (log2 TPM) by Tarlatamab response (p-value, t-test, error bars 95% CI) (B) Receiver-operater curve for Tarlatamab response prediction with DLL3 expression (log2 TPM). (C) Distribution of SCLC transcription factor subtype assigned by transcriptomic signature classification by Tarlatamab response.

In this 12-patient cohort, a log₂ *DLL3* TPM cutoff of 3.8 provided the strongest discrimination between responders and non-responders, achieving an AUC of 0.83 for predicting tarlatamab response (sensitivity 0.833, specificity 0.833, Figure 2B). We benchmarked this cutoff using a large external RNA-seq cohort of 307 publicly available SCLC samples, and 39.3% of tumors exceeded this *DLL3* expression threshold (Figure S2).

Overall, *DLL3* expression determined by clinical whole-transcriptome sequencing was predictive of tarlatamab response and corresponded to a tumor fraction similar to the observed response rate in the phase 2 DeLLphi-301 trial, which supports its potential as a practical biomarker for clinical stratification.

### ASCL1-driven transcriptional subtype is enriched among tarlatamab responders

Given the emerging importance of transcriptional subtypes in SCLC biology and therapeutic response, we applied a transcriptional subtype classifier to assign each sample to a canonical molecular subtype using established gene expression signatures (adapted from (Park et al. 2024), Figure S3A). Samples were sorted into five subtypes: SCLC-A (ASCL1-driven), SCLC-N (NEUROD1-driven), SCLC-A/N (mixed), SCLC-P (POU2F3-driven, tuft-cell–like), and SCLC-TN (triple-negative), using a hierarchical supervised classifier applied to the full RNA-seq cohort (n = 321, Figure S3B).

Applying this classifier to the full 321 sample cohort reproduced the expected subtype distribution (39.9% SCLC-A, 31.9% SCLC-A/N, 18.4% SCLC-N, 4.1% SCLC-P, and 5.9% SCLC-TN, Figure S3C). ASCL1, NEUROD1, and Tuft-cell gene signatures showed the expected specificity to sorted transcriptional subtypes (Figure S3D). *DLL3* expression was highest in SCLC-A and A/N tumors (Figure S4A), and SCLC-TN tumors were differentiated by expression of *YAP1* (Figure S4B).

Within the tarlatamab-treated subset (n = 12), most tumors were SCLC-A (7/12), followed by SCLC-A/N (3/12) and SCLC-N (2/12, Figure S3C). Among responders, the majority were SCLC-A (5/6) with 1/6 classified as SCLC-A/N (Figure 2C). In contrast, non-responders were distributed as 2/6 SCLC-N, 2/6 SCLC-A, and 2/6 SCLC-A/N, a difference that was not statistically significant given the limited sample size (p = 0.1637, chi-square test, Figure 2C).

When viewed through the lens of transcription factor subtypes, tarlatamab responders appear to be predominantly ASCL1-driven, while NEUROD1-driven (SCLC-N) tumors were exclusively non-responders, suggesting potential subtype-specific biological differences in DLL3 dependency and immune microenvironment.

### Inflamed tumor immune microenvironment and *DLL3* expression jointly define tarlatamab response

Having identified that tarlatamab-responsive tumors exhibit high *DLL3* expression and enrichment for the ASCL1 transcriptional subtype, we next examined whether features of the tumor immune microenvironment further distinguished responders from nonresponders. Prior studies have shown that SCLC immune phenotypes influence response to immunotherapy^5,16^ prompting us to test whether inflammation-related transcriptional programs were associated with tarlatamab sensitivity.

TIL counts and ratios were collected for 11 pretreatment and 2 posttreatment biopsies and there were no significant differences between responders and nonresponders (mean counts 11.8 vs 15.4; p=0.4444; mean ratio 4.89×10^−5^ vs 6.61×10^−5^; p=0.4177, t-test). Unsupervised differential expression testing of responder versus nonresponder tumors revealed strong enrichment of immune-related pathways in responders, including leukocyte-mediated immunity, cytotoxicity, chemotaxis, and cell-cell adhesion, suggesting a more inflamed tumor microenvironment (Figure S5). The sole pathway enriched in nonresponders was related to neuronal differentiation, likely driven by the presence of SCLC-N tumors in this group.

Using a supervised approach, we calculated three established immune gene signatures in the full SCLC whole-transcriptome cohort (n=321, NK cell, T cell inflamed, and cytotoxic T cell; see Methods) and averaged them to derive a composite immune activity score (Figure S3A). Tarlatamab-responsive tumors (n=6) clustered toward the most inflamed end of this spectrum, whereas nonresponders showed lower immune activity (mean –0.08 vs –0.27; p = 0.061, t-test). When placed in the context of the larger SCLC cohort, responders mapped to the upper range of immune activity (median 67th percentile, range 10 to 99), while nonresponders fell much lower (median 4th percentile, range 0.3 to 64). One notable exception was patient 10, a nonresponder whose high immune activity (64th percentile) contrasted with its SCLC-N transcriptional subtype and minimal *DLL3* expression (Figure S3A). This contrast highlights the interaction between *DLL3* expression, transcriptional subtype, and the tumor immune microenvironment. To illustrate the combined predictive potential of *DLL3* expression and immune activity, we calculated a composite *DLL3* x Immune Activity score (DLI score, see Methods) which further separated responders and non-responders (mean composite score 0.723 vs −0.713, p = 0.013, t-test, sensitivity 0.83, specificity 1, Figure 3B).

**Figure 3.**
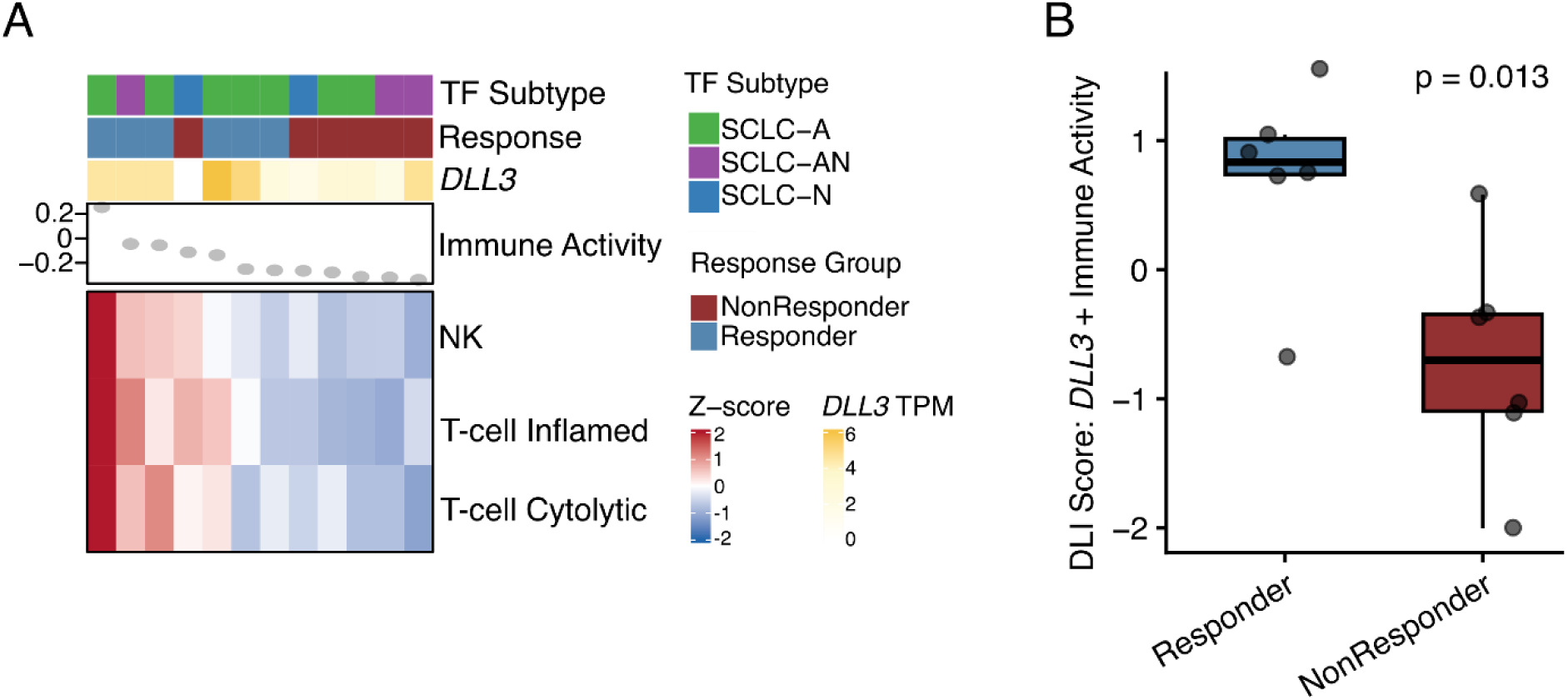
(A) Summary of immune microenvironment of treated tarlatamab tumors through NK, cytolytic T cell, T cell inflamed, and aggregate immune activity score by clinical response, TF subtype, *DLL3* expression. (B) Composite *DLL3* expression and immune activity (DLI) score by response to tarlatamab.

**Figure 4.**
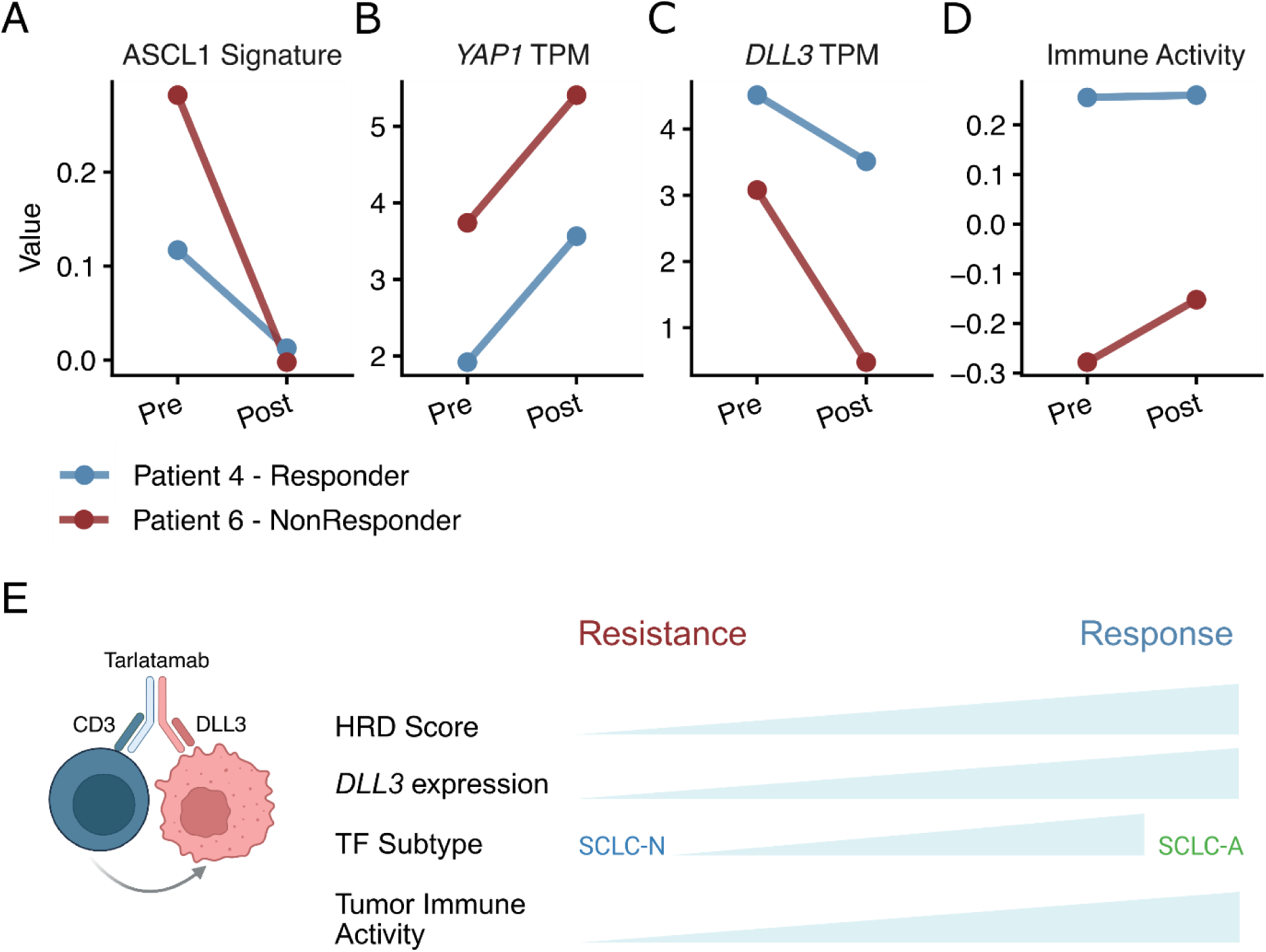
Phenotypic changes in post-progression samples, including (A) ASCL1 transcriptional signature score (singscore mean), (B) *YAP1* expression (TPM), (C) *DLL3* expression (TPM), and (D) immune activity score (singscore mean). (E) Integrated model for molecular features of SCLC Tarlatamab response.

Together, these findings support a model in which *DLL3* expression, ASCL1-driven transcriptional state, and an inflamed tumor immune microenvironment jointly characterize tumors most likely to respond to tarlatamab.

### Progressive tumors on tarlatamab develop *MYC*-family alterations, lose ASCL1 identity, downregulate *DLL3*, and gain YAP1 expression

We next examined whether the molecular features associated with tarlatamab response changed following disease progression. Two patients, one with an inflamed phenotype who initially achieved a partial response (Patient 4) and one with an immune-cold phenotype with primary progression (Patient 6), underwent post-progression biopsies analyzed by whole-exome and whole-transcriptome sequencing. Immunohistochemistry revealed a loss of synaptophysin in Patient 4’s post-treatment biopsy. Ki67 proliferation index was 80% pre-treatment and 95% in Patient 4’s post-treatment biopsies.

Across both paired pre- and post-tarlatamab samples, core lineage-defining SCLC drivers such as TP53/RB1 loss, and *KMT2D* mutation remained stable, confirming shared tumor origin (table S2). However, both patients demonstrated clonal evolution at progression, including the emergence of MYC-family activation (*MYCN* mutation and *MYCL* amplification), expansion of *NOTCH2* and *NOTCH3* alterations, and increased structural variation affecting genes such as *LRP1B* and *RB1* (table S2). These patterns collectively suggest that tarlatamab resistance is accompanied by *MYC*-driven transcriptional reprogramming, NOTCH pathway dysregulation, and increased genomic instability.

Both tumors showed convergent transcriptional evolution after progression. Although both initially exhibited ASCL1-high, SCLC-A phenotypes, their post-progression tumors showed a marked loss of ASCL1 signature expression compared to the full SCLC cohort, (Patient 4: 34th to 8th percentile; Patient 6: 99th to 7th percentile) and were classified as SCLC-TN, consistent with lineage plasticity and loss of neuroendocrine differentiation. In parallel, *YAP1* expression increased substantially (Patient 4: 38th to 73rd percentile; Patient 6: 76th to 97th percentile), suggesting a shift toward a YAP1-driven, non-neuroendocrine state.

As predicted by this lineage change, *DLL3* expression was markedly reduced following progression (Patient 4: 4.51 to 3.51 TPM, 81st to 51st percentile; Patient 6: 3.08 to 0.48 TPM, 40th to 10th). In contrast, the composite immune activity score remained stably high in the responder (99th to 100th percentile) but showed an increase in the nonresponder (5th to 48th percentile.

Together, these findings indicate that progression after tarlatamab is associated with clonal evolution and *MYC* alterations, de-differentiation from an ASCL1-driven, DLL3-high neuroendocrine state toward a YAP1-driven, transcriptionally plastic phenotype, consistent with lineage escape mechanisms described in SCLC progression.

## Discussion

SCLC outcomes remain poor despite advances in therapy. Tarlatamab has shown meaningful activity in relapsed SCLC in the phase II DeLLphi-301 trial, but biomarkers for response prediction and patient selection remain largely unknown. In this study, we leverage integrated whole-exome and whole-transcriptome sequencing to identify molecular correlates of tarlatamab response in 12 SCLC patients. The clinical activity of tarlatamab was consistent between this cohort and the DeLLphi trials, providing a representative context to examine tumor-intrinsic and immune correlates of therapeutic sensitivity. Through integrative analysis, we identified associations involving *DLL3* expression, transcriptional lineage, and enrichment of immune-related pathways, suggesting that response to tarlatamab reflects the interaction of tumor identity, target expression, and immune contexture. Although somatic driver alterations did not differ by response, as expected given the genetic homogeneity of SCLC, homologous recombination deficiency scores were higher in responsive tumors, raising the possibility that increased genomic instability may modulate sensitivity to tarlatamab.

The role of *DLL3* expression in tumor response to tarlatamab is currently understudied. Here, we identify an association between increased levels of expression with response to tarlatamab. Using a log₂ *DLL3* TPM cutoff of 3.8 benchmarked using a large external RNA-seq cohort, we achieve an AUC of 0.83 to predict tarlatamab response. This is consistent with the DLL3 based mechanism of action and highlights that RNA-based assessment of *DLL3* expression may further discriminate tumors responsive and non-responsive to tarlatamab. This is consistent with similar recent findings using single circulating tumor cell protein staining of DLL3^32^. An association between *DLL3* expression and tumor response has implications for integration with other therapies^33^ as well as treatment of other *DLL3* expressing tumors^34^.

Interpretation of *DLL3* expression and response is further contextualized by transcriptional lineage. ASCL1-driven tumors were more likely to respond to tarlatamab and were associated with higher *DLL3* expression and a neuroendocrine phenotype, whereas NEUROD1- and YAP1-driven driven tumors were more resistant to tarlatamab, and were associated with lower *DLL3* expression and features of immune exclusion. Analysis of post-tarlatamab tumors further revealed a shift from ASCL1 toward YAP1 expression, suggesting transcriptional plasticity and lineage escape as potential mechanisms of acquired resistance. These transitions mirror evolutionary trajectories previously described following chemotherapy and immune checkpoint blockade^4,5^, indicating that lineage reprogramming may represent a shared resistance pathway across SCLC treatment modalities. Notably, prior work has described a MYC-independent incompatibility between ASCL1 and NEUROD1 co-expression^35^, suggesting biological constraints on certain lineage transitions. Consistent with this framework, we did not observe emergent ASCL1/NEUROD1 co-expression in our cohort, supporting a model in which resistance is mediated by shifts toward alternative lineage states, such as YAP1, rather than unrestricted plasticity across neuroendocrine programs.

Finally, increased expression of cytotoxic and leukocyte-mediated immune pathways was associated with response to tarlatamab, a finding that is consistent with its mechanism of action as a bispecific T-cell engager requiring functional immune effector activity. Emerging evidence supports the presence of distinct immune subtypes in SCLC, with differential responses to chemotherapy^4^ and immune-based therapies^4,5^, underscoring the biological relevance of immune context in this disease. In this setting, our findings suggest that *DLL3* expression alone may be insufficient to drive therapeutic activity in the absence of pre-existing or inducible immune activation. Primary resistance appears to reflect an unresponsive baseline lineage and immune context, while acquired resistance is associated with dynamic transcriptional reprogramming and lower *DLL3* expression. Notably, TIL counts and ratios did not distinguish responders from non-responders, which emphasizes functional immune activity over immune cell presence. γδ T-cell–mediated cytotoxicity has been associated with improved response to anti–PD-L1 therapy and contribution in tarlatamab-mediated tumor cell killing which further supports the importance of immune functional state in response modulation^36^.

This study is limited by its relatively small, single-institution cohort. Despite this limitation, we outline a biological model in which response to tarlatamab is associated with the integration of target expression, lineage context, and immune state. Our findings further implicate lineage reprogramming as a potential mechanism of acquired resistance that can be evaluated using integrated clinical WES and WTS profiling. Prospective studies^37^ will be necessary to validate these observations, define mechanisms of response and resistance, and enhance their clinical utility.

## Supplemental Material

**Table S1.** Sequencing Quality Metrics for Whole-Exome Sequencing (WES) and Whole-Transcriptome Sequencing (WTS). Note: WES samples N=14; WTS samples N=14.

| Platform | Metric | Mean | Median | SD | Min | Max |
| --- | --- | --- | --- | --- | --- | --- |
| WES | Mean Target Coverage (X) | 282.2 | 277.6 | 67.1 | 148.0 | 419.2 |
| | % Target Bases $\geq 20\times$ | 97.0% | 97.0% | 0.0% | 97.0% | 97.0% |
|  | % Bases On-Target | 25.7% | 26.0% | 4.1% | 14.0% | 31.0% |
|  | Duplicate Rate | 15.6% | 15.0% | 5.10% | 8.0% | 30.0% |
|  | FOLD-80 Base Penalty | 1.84 | 1.85 | 0.13 | 1.66 | 2.06 |
| WTS | Sequencing Depth (Gb) | 22.0 | 19.2 | 8.32 | 12.9 | 42.0 |
|  | % Aligned Bases | 96.4% | 96.6% | 0.91% | 94.3% | 97.3% |
|  | % Coding Bases | 76.0% | 75.8% | 1.79% | 73.3% | 79.8% |
|  | % Ribosomal Bases | 0.51% | 0.50% | 0.39% | 0.06% | 1.24% |
|  | Duplicate Rate | 51.1% | 53.0% | 8.35% | 32.0% | 64.0% |

**Figure S1.**
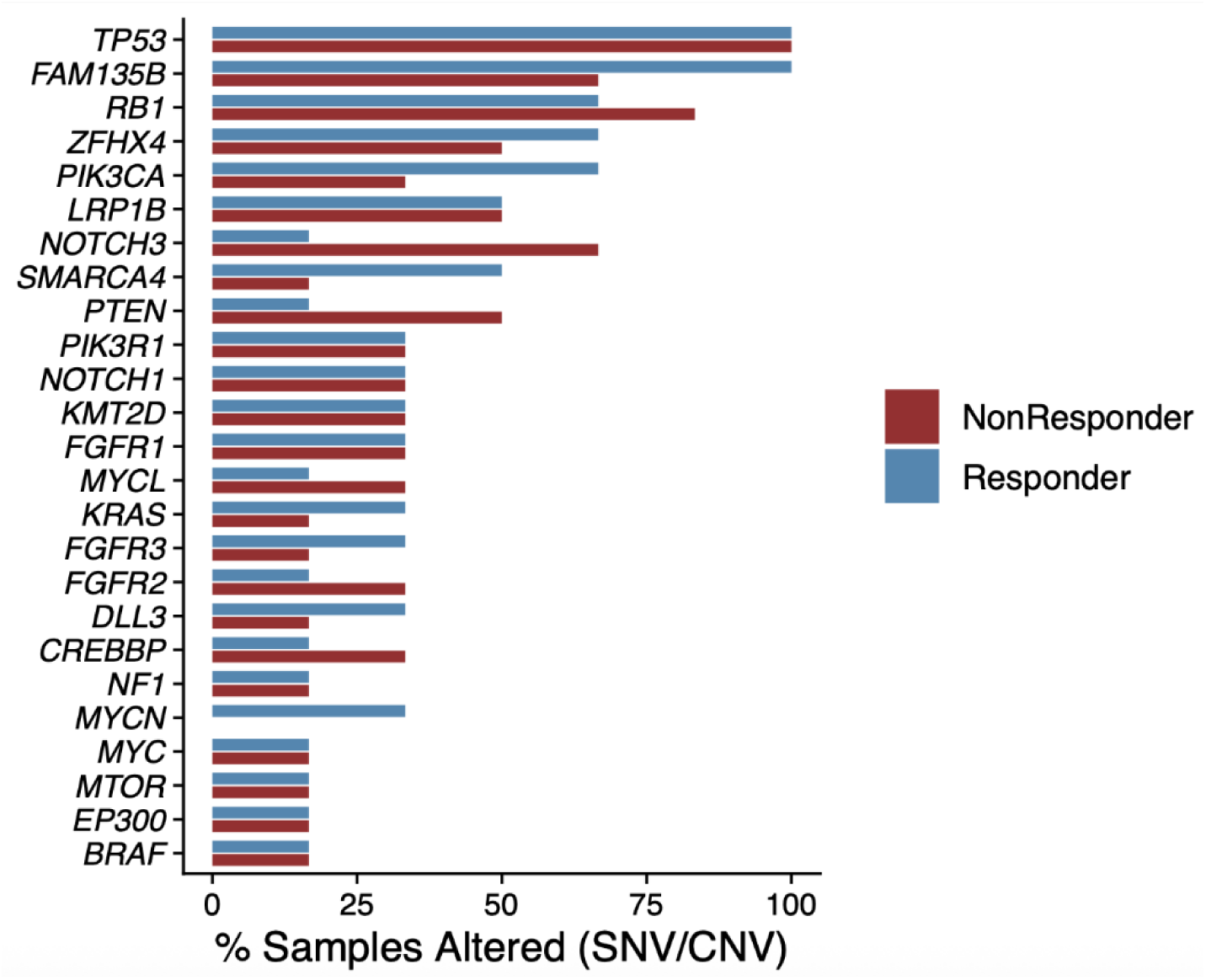
Percentage samples with somatic or copy number alterations in key recurrent SCLC driver genes.

**Figure S2.**
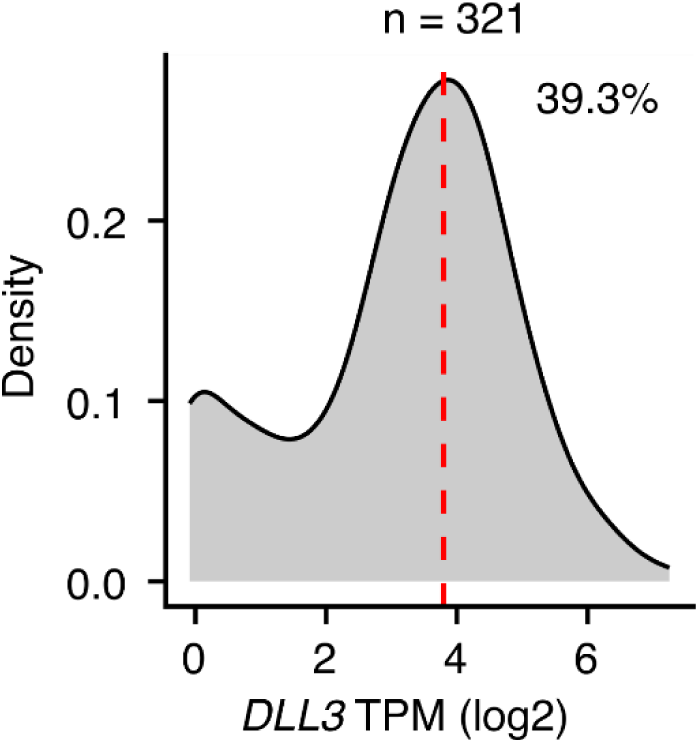
Distribution of *DLL3* expression (log2 TPM) across full SCLC cohort (n = 321), red dotted line TPM = 3.8, proposed discriminatory cutoff for tarlatamab response.

**Figure S3.**
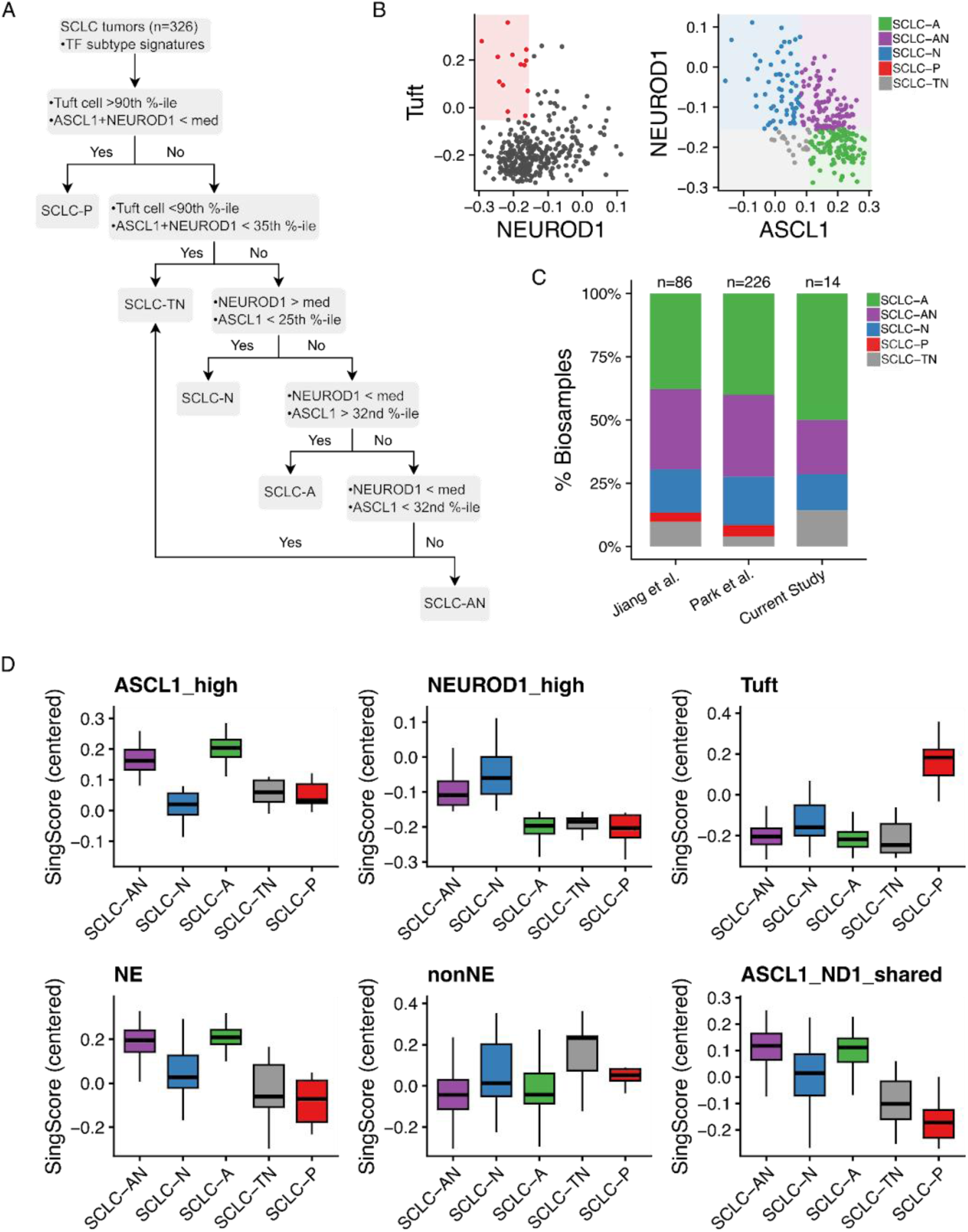
(A) Sorting schema for SCLC transcriptomic TF subtype classifier. (B) Tuft, NEUROD1, ASCL1 signature expression (singscore) enables TF subtype sorting. (C) Distribution of transcriptomic TF subtype by included study. (D) Signature expression (singscore) by sorted samples in full cohort (n=321, NE = neuroendocrine, nonNE = non neuroendocrine, ND1 = NEUROD1).

**Figure S4.**
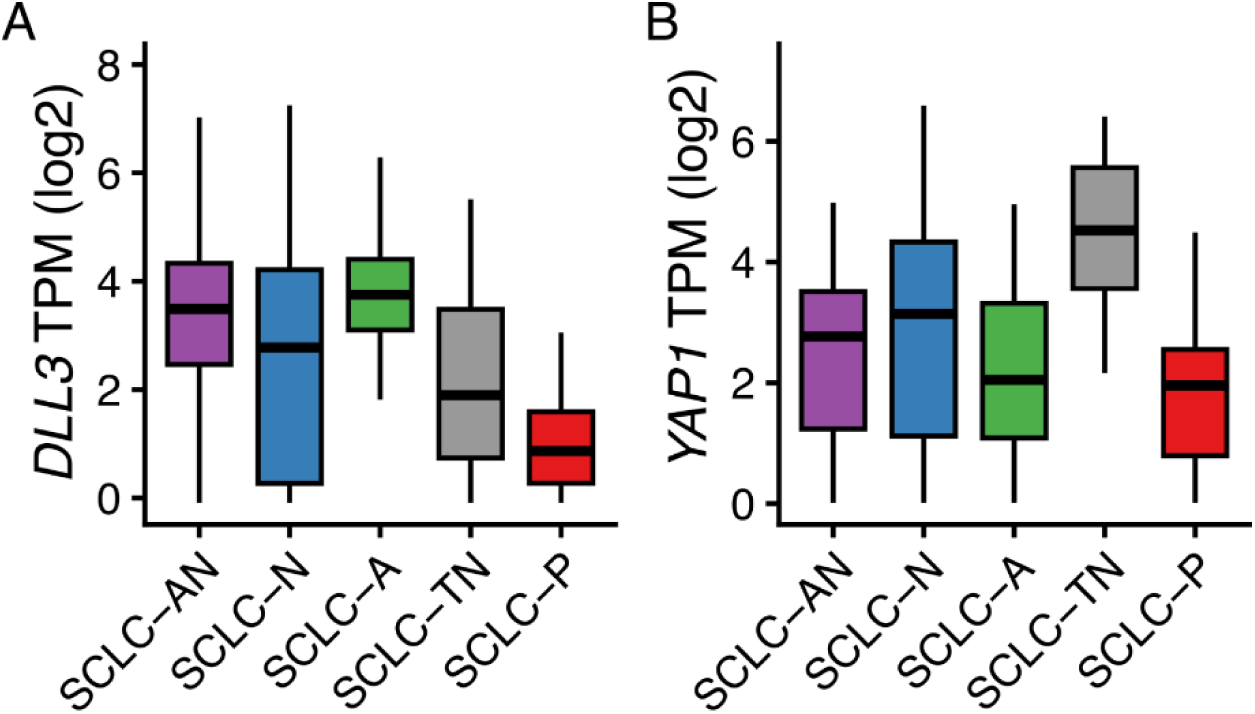
(A) *DLL3* and (B) *YAP1* expression (log2 TPM) across full cohort (n=321) by sorted TF subtype.

**Figure S5.**
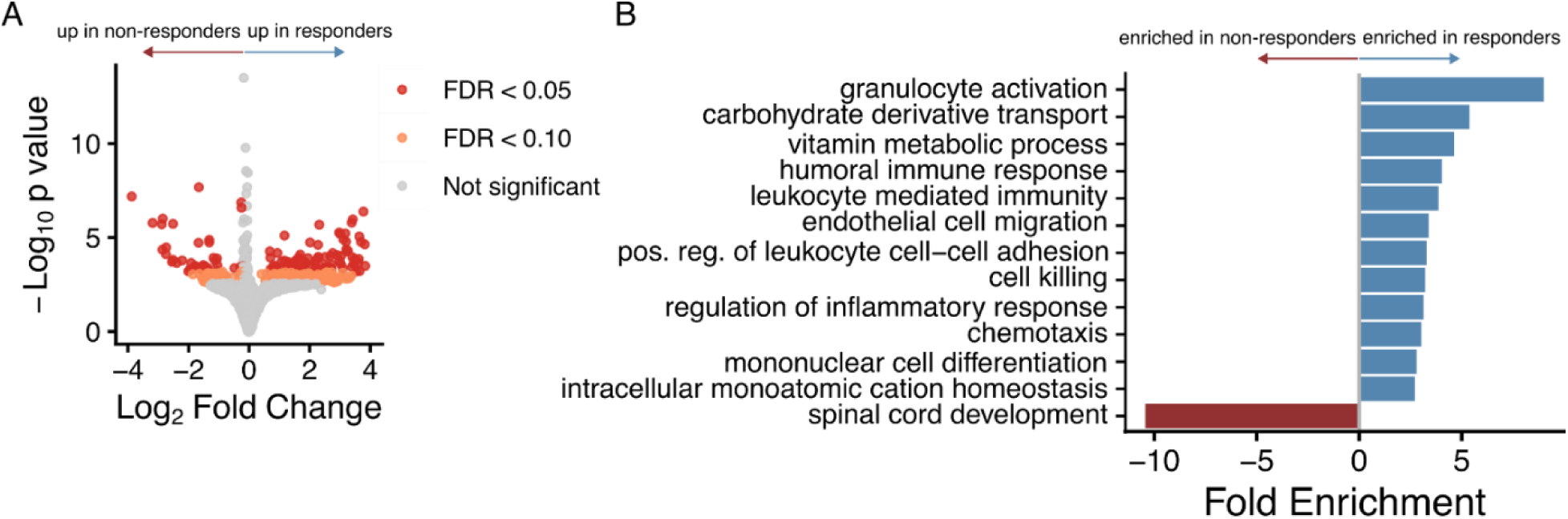
(A) Volcano plot and (B) gene set enrichment analysis of differential expression in tarlatamab responders (n = 6), versus nonresponders (n = 6) at FDR < 0.05 and <0.10).

**Table S2.** Key genomic alterations in paired pre- and post-progression samples. Legend: ✓ = alteration present, — = alteration absent. Log2 copy number ratio in parentheses for CNV.

| Gene (alteration) | Pt 4 pre | Pt 4 post | Pt 6 pre | Pt 6 post |
| --- | --- | --- | --- | --- |
| *MYCL* (amplification) | — | ✓ (0.90) | — | ✓ (3.1) |
| *MYCN*<br>(missense) | — | ✓ | — | ✓ |
| *NOTCH2* (mutation) | ✓ | ✓ | ✓ | ✓ |
| *NOTCH3*<br>(missense) | — | — | ✓ | ✓ |
| *LRP1B*<br>(missense) | — | — | — | ✓ |
| *LRP1B*<br>(deletion/missense) | ✓(-1.9) | ✓(-3.3) | — | ✓ |
| *RB1* (loss/frameshift) | ✓ | ✓ | ✓ | ✓ |
| *TP53* (mutation) | ✓ | ✓ | ✓ | ✓ |
| *NF1* (missense) | — | — | — | ✓ |
| *DLL3* (amplification) | — | — | — | ✓(0.93) |
| *KMT2D* (mutation) | — | — | ✓ | ✓ |
| *PIK3CA*<br>(amplification) | — | — | ✓ (0.76) | ✓ (0.95) |

